# Routine Saliva Testing for SARS-CoV-2 in Children: Partnering with Childcare Centers in the Greater New Haven Community

**DOI:** 10.1101/2022.05.05.22274434

**Authors:** Erica J. Rayack, Hibah Mahwish Askari, Elissa Zirinsky, Sarah Lapidus, Hassan Sheikha, Chikondi Peno, Yasaman Kazemi, Devyn Yolda-Carr, Chen Liu, Nathan D. Grubaugh, Albert I. Ko, Anne L. Wyllie, Erica S. Spatz, Carlos R. Oliveira, Amy K. Bei

## Abstract

**Background:** While considerable attention was placed on SARS-CoV-2 testing and surveillance programs in the K-12 setting, younger age groups in childcare centers were largely overlooked. Childcare facilities are vital to communities, allowing parents/guardians to remain at work and providing safe environments for both children and staff. Therefore, early in the COVID-19 pandemic, we established a PCR-based COVID-19 surveillance program in childcare facilities, testing children and staff with the goal of collecting actionable public health data and aiding communities in the progressive resumption of standard operations and ways of life. In this study we describe the development of a weekly saliva testing program and provide early results from our experience implementing this in childcare centers.

**Methods:** We enrolled children (aged 6 months to 7 years) and staff at 8 childcare facilities and trained participants in saliva collection using video chat technology. Weekly surveys were sent out to assess exposures, symptoms, and vaccination status changes. Participants submitted weekly saliva samples at school. Samples were transported to a partnering clinical laboratory for RT-PCR testing using SalivaDirect and results were uploaded to each participant’s online patient portal within 24 hours.

**Results:** This study fostered a cooperative partnership with participating childcare centers, parents/guardians, and staff with the goal of mitigating COVID-19 transmission in childcare centers. Age-related challenges in saliva collection were overcome by working with parents/guardians to conceptualize new collection strategies and by offering parents/guardians continued virtual guidance and support.

**Conclusion:** SARS-CoV-2 screening and routine testing programs have focused less on the childcare population, resulting in knowledge gaps in this critical age group, especially as many are still ineligible for vaccination. SalivaDirect testing for SARS-CoV-2 provides a feasible method of asymptomatic screening and symptomatic testing for children and childcare center staff. Given the relative aversion to nasal swabs in the childcare age group, especially as a routine surveillance tool, an at-home saliva collection method provides an attractive alternative. Results can be shared rapidly electronically through participants’ private medical chart portals, and video chat technology allows for discussion and instruction between investigators and participants.

## Background

As SARS-CoV-2 transmission continues, vaccination efforts proceed, and communities reopen, surveillance remains a valuable tool for mitigating the effects of COVID-19. With children under 5 years of age currently ineligible for vaccines available in the United States, screening for SARS-CoV-2 stands as a vital control strategy, with ongoing research into the acceptability and effectiveness of screening methods remaining an integral component. COVID-19 screening programs are of particular importance in childcare centers [1] as they could identify infections early, prevent outbreaks, and keep centers open, particularly in times of high community viral transmission.

The use of saliva samples for surveillance of SARS-CoV-2 infections in children opens doors to increased feasibility and reduced invasiveness in testing across a wide range of ages. SalivaDirect was granted Emergency Use Authorization from the U.S. Food and Drug Administration in August 2020 and showed greater sensitivity to SARS-CoV-2 than nasopharyngeal sampling, with low rates of false-positive and invalid results [2]. This simple, effective, and non-invasive testing method provides an alternative to methods that require expensive additives and costly cooling approaches, making it suitable for the needs of large-scale testing [3].

Design of this study focused on strong partnerships with families and childcare centers directors and staff to inform study execution. The goals were to mitigate outbreaks in daycare centers and nursery schools, provide actionable public health data, and aid communities in the progressive resumption of standard operations and ways of life by identifying cases to prevent transmission. The following is a guide to establishing a surveillance program in childcare facilities using SARS-CoV-2 saliva RT-PCR testing in children and staff.

## Methods/Design

### Recruitment and Enrollment

We enrolled children between the ages of 6 weeks to 7 years and staff of 7 childcare facilities. To begin, we organized town-halls and small discussions with parents/guardians and staff of participating childcare facilities via Zoom. The study team described the purpose, goals, and design of the study, and staff and parents/guardians were given an opportunity to voice concerns and propose suggestions. We discussed the timing of saliva collection, teaching/observation method for collection (by video), and notification and timing of test results. The study design was iteratively updated with considerations for both staff and parent suggestions (in addition to IRB and health and safety requirements), recognizing the importance of partnering with childcare centers and increasing the likelihood of stakeholder buy-in and compliance within each center. The goal was to make this process easy for families and staff of these centers in order to ultimately improve the coverage and timeliness of testing results. Consent documentation was emailed or dropped off at childcare facilities for parents/guardians and staff to pick up at their convenience. Once a prospective participant had the consent document, a virtual meeting was scheduled for each participant to review and sign the consent, provide basic demographic information, and complete the initial saliva collection with a study team member.

### Saliva Sample Collection

Participants received weekly collection materials including empty saliva collection tubes, disposable plastic bulb transfer pipettes, tube labels, and biohazard bags. These supplies were available for participants to pick up in designated coolers outside of school at the drop-off location each week. Initial saliva collections were conducted virtually over Zoom, with study staff guiding participants (parents/guardians, children, and staff). Participants were provided with conical collection tubes approved by the testing facility (ranging in size from 5ml-50 mL) and provided 0.5-1 mL of saliva. Saliva collection techniques varied by age, as outlined below. Participants collected samples at home in the evening, refrigerated them overnight, and dropped them off the following morning in a designated cooler outside of each center. Figure 1 provides detailed instructions for saliva collection by age groups.

**Figure 1.**
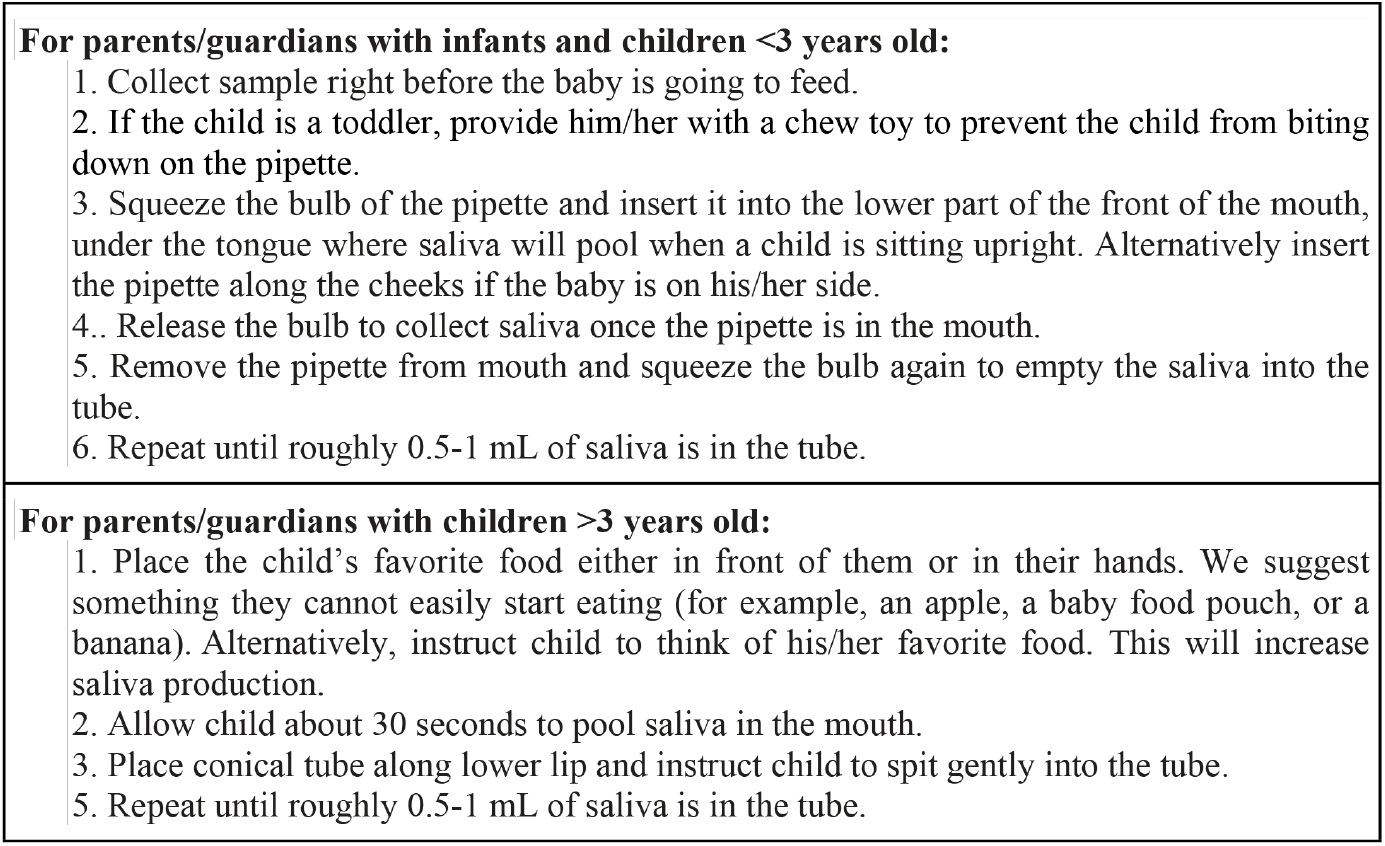
Instructions for Saliva Collection.

Participants were considered fully trained in collection after 2-3 virtually supervised sessions and, after reporting no difficulty, could perform the subsequent collections unsupervised. If parents/guardians or staff had challenges following this, additional sessions were scheduled. The study team would have an open “drop-in” Zoom link with a waiting room where families could join one by one the evening before scheduled drop-offs should they require additional assistance or coaching. Participants received a weekly email to remind them to pick up collection materials, collect saliva samples at home, and drop them off at their respective childcare facilities the following morning. Participants were instructed to label their tube and place it in the biohazard bag for storage overnight in their refrigerator (or collect it in the morning before school). Designated coolers were provided to the childcare facilities by the study team to facilitate the sample drop-off, and liaisons at the childcare facilities were reminded to place the coolers in the drop-off area for parents/guardians.

### Transportation, Storage, and Processing

Following participant sample drop-off, coolers were collected by a designated study team member with approvals for transporting samples. They were transported to the Yale Pathology laboratory, where they were processed in a CLIA-approved laboratory within 24 hours using the SalivaDirect RT-PCR assay to detect SARS-CoV-2 RNA.

Participants were given access to the Yale New Haven Hospital (YNHH) MyChart platform to view test results. Participants were not contacted in the event of a negative result unless specifically requested by the participant. Positive results were directly reported to the participant and reported to the study physician. The study physician then contacted the participant to provide guidance and instruction on isolation and continuity of care with the participant’s primary care provider as needed. Parents/guardians and staff consented to notify their respective childcare facility in the case of a positive result. Childcare facility directors were also notified if no positives were detected that week, allowing for the continuation of routine operations.

### Data Collection

Research Electronic Data Capture (REDCap) software was used to record study data. Weekly surveys were also sent out using this platform. The survey allowed for the tracking of symptoms, recent travel and exposure histories, SARS-CoV-2 vaccination status, and the number of days of childcare attended per week.

## Results and Discussion

### Participation and Acceptability

Age-related challenges in sample collection were overcome through engagement and discussion with parents/guardians and their children. Guidance in the form of written instructions, visual aids, and optional weekly video chatting with study staff provided parents/guardians with the support needed to improve saliva collection feasibility for both infants and toddlers. Frequent email communications sent to participants emphasized the availability of study staff to help with collection or attend to study-related concerns. Notably, when reminder emails were not sent to parents/guardians and facility liaisons, participation for that specific week dropped dramatically, again elucidating the importance of frequent engagement between the study team and parents/guardians. Key challenges and potential remedies are summarized in Figure 2.

**Figure 2.**
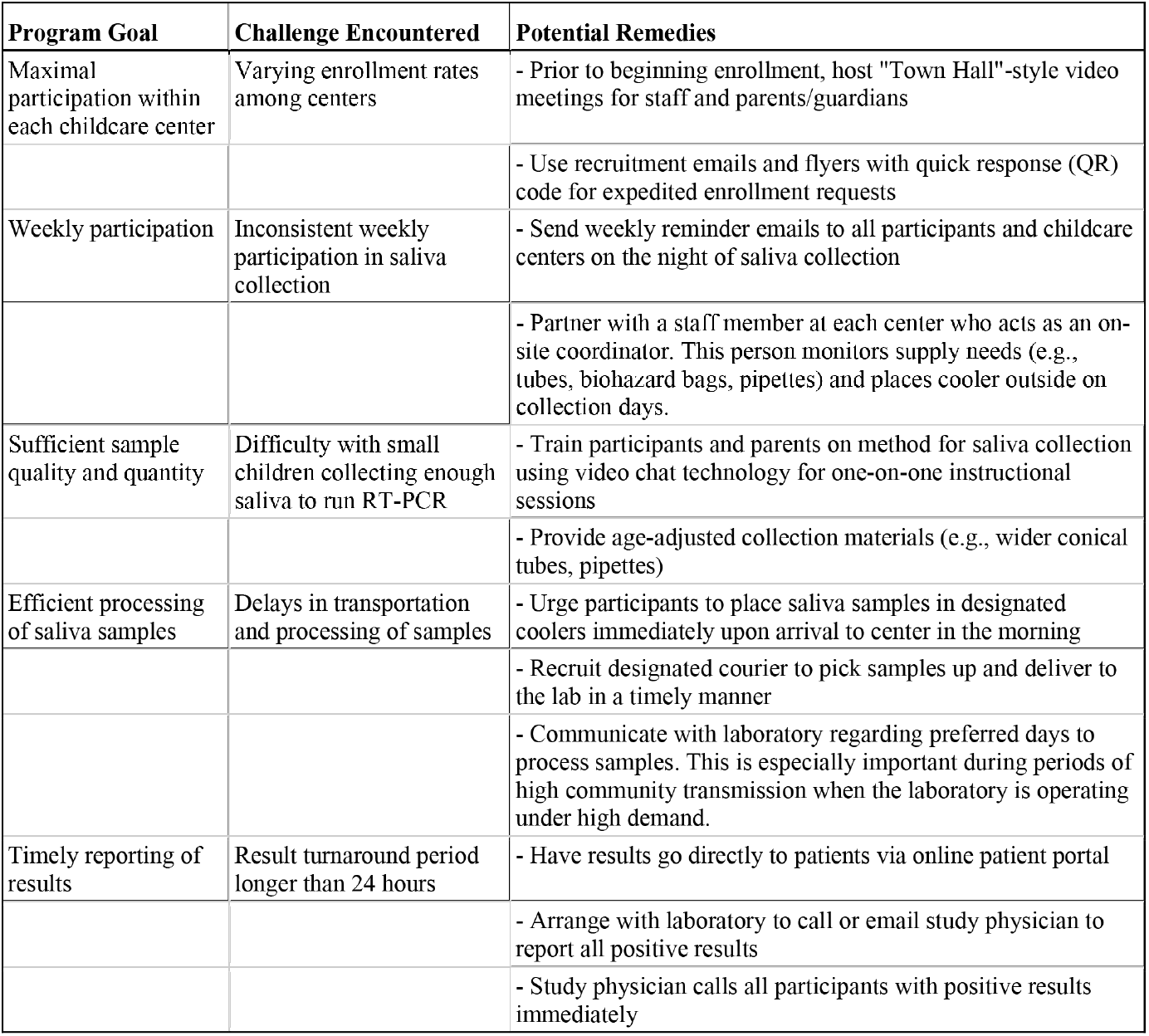
Challenges and Potential Remedies of Weekly Saliva Testing in Childcare Centers.

A considerable drop in inconclusive samples was observed as the study proceeded. Communication with the laboratory staff revealed insufficient volume or tube leakage as the main causes of inconclusive samples. Study materials were adjusted to increase the ease of tube closure, and reminder emails were sent to participants regarding the minimal saliva volume needed and the importance of tightening tube lids. The ability of participants to adapt to these instructions supports the feasibility of at-home saliva sample collection.

Despite the initial transient challenges to at-home saliva collection, it offers a less-invasive alternative to nasal/nasopharyngeal swabs in young children, especially when accompanied by written instruction for parents/guardians. Collection completion by parents/guardians and their children decreases the need for interaction with the healthcare workforce, thereby decreasing the risk of nosocomial infection and alleviating a major factor in testing bottlenecks [4, 5]. It also alleviates the need for supplies, such as nasal swabs and personal protective equipment [4].

#### Frequency of Testing

The public health value of frequent asymptomatic SARS-CoV-2 testing has been emphasized in multiple analytic modeling studies [6, 7]. Models have demonstrated that sporadic testing is likely to increase missed positive tests or first-time positive tests from individuals who have already passed their infectious period [7]. Weekly testing was sufficient to provide effective attenuation of infection surges in models, while less frequent testing, or no testing, was not [7]. Prompt reporting of results was also highly supported by these models [7]. Based on the information presented in these models, weekly surveillance with next-day reporting of test results was implemented in this study.

Though more frequent testing is ideal for infection monitoring, our study was designed to balance effectiveness, feasibility, and the unique circumstances occurring at this time. For instance, Environmental Health and Safety protocols were frequently adjusting and updating in response to the changing state of the pandemic, resulting in our further reliance on tools, such as video communication and email discussion as opposed to face-to-face approaches. Our methods can be practicably adapted to contexts beyond childcare facilities. Procedural adaptation must account for variations by context, including transmission dynamics, cost-effectiveness, and community features [6, 7].

## Conclusions

Childcare centers are critical to communities and provide services essential to society. Weekly screening for SARS-CoV-2 infections among children and staff may mitigate outbreaks and allow centers to remain open more consistently. Here, we conducted weekly screening using SalivaDirect RT-PCR. At-home saliva testing is a simple and noninvasive alternative to nasopharyngeal swabs and is optimally suited for routine and frequent testing for surveillance purposes outside a hospital setting. In partnership with childcare centers and parents/guardians, we were able to implement routine saliva-based testing for SARS-CoV-2 with ease in this setting. Weekly SalivaDirect testing in childcare centers will continue to prove beneficial as new variants continue to emerge and community rates fluctuate.

## Data Availability

All data produced in the present work, namely the details of the study protocol, are contained in the manuscript.

## Ethical Statement

Ethical approval for this study was granted by the Human Investigation Committee of Yale University (Protocol 2000028639). All samples were collected with informed consent and in accordance with all ethical requirements of the Human Investigation Committee of Yale University.

## Acknowledgements

We would like to acknowledge Beth Bye, Commissioner for the Connecticut Office of Early Childhood (OEC) and Rachel Leventhal-Weiner at the Connecticut Office of Early Childhood (OEC) for useful discussions on the strategy and impact of the study. We would also like to thank the directors, staff, and families of the childcare centers in our study: Westville Community Nursery School, Alphabet Academy and Nest Schools, Friends Center for Children – Blake Street, Friends Center for Children – E. Grand, and the Jewish Community Center of Greater New Haven. We would like to thank useful discussions with Dr. Harlan Krumholz on the vision and strategy of the approach.

